# Implementation and impact of a 5-year community-based tuberculosis screening intervention in Cambodia: a mixed-methods pragmatic evaluation using the RE-AIM framework

**DOI:** 10.64898/2026.05.29.26354425

**Authors:** Brenda Soun, Chamroen Pall, Michiko Nagashima-Hayashi, Hsandy Thovy, Saren Menh, Seyha Ong, Sovanvorleak Tep, Sothearith Eng, Khay Mar Aung, Siyan Yi, Sok Chamreun Choub, Sovannary Tuot, Alvin Kuo Jing Teo

## Abstract

**Background:** Cambodia is a high-TB burden country where over a third of TB cases have gone undetected. The Community Mobilisation Initiatives to End TB (COMMIT) programme, implemented across four provinces and 27 operational districts (ODs) in Cambodia from October 2019 to September 2024, aimed to improve TB case finding, diagnosis, treatment, and prevention through community-driven approaches. This study evaluated the implementation, programme outcomes, and sustainability of COMMIT to inform future TB initiatives.

**Methods:** This mixed-methods explanatory sequential study used the Reach, Effectiveness, Adoption, Implementation, and Maintenance (RE-AIM) framework. Quantitative data were collected from the programme database and the national TB Management Information System (TB-MIS). In-depth interviews, guided by the Theoretical Domains Framework (TDF), explored contextual factors influencing programme implementation and complement quantitative findings. Quantitative data were analysed descriptively to estimate screening coverage, diagnostic yield, and construct care cascades. Qualitative data were transcribed and translated into English, coded, consolidated into a matrix structured using RE-AIM and TDF components, and analysed thematically.

**Results:** COMMIT screened 695,970 people for TB. Key populations were reached, though sex and age disparities in screening participation reflected underlying social and structural barriers. Approximately 98% of those screened underwent diagnostic testing. Treatment initiation (>99%) and completion (>97%) rates were high. COMMIT operationalised contact investigation and evaluation for TB preventive treatment (TPT), screening over 90% of notified contacts. More than 20,000 people were TPT-eligible, of whom 68.7% enrolled in and 86.2% completed TPT. These programme outcomes were supported by strong community engagement, expansion of rapid molecular diagnostics, and programme adaptability during COVID-19. COMMIT was scaled from 10 to 27 ODs, during which it strengthened community capacity by training healthcare workers and expanding peer support groups. Stakeholders emphasised the need to reinforce local ownership and public-private sector collaboration, strengthen integrated services, and de-implement low-value practices such as symptom-based screening.

**Conclusions:** COMMIT improved TB case detection, treatment support, and prevention in Cambodia through community-led strategies and sustained capacity building. Maintaining the programme’s impact will require continued investment in community systems, de-implementation of low-value practices, and the adoption of efficient, person-centred approaches that address evolving community needs.

## Introduction

Tuberculosis (TB) is the leading cause of morbidity and death from infectious diseases globally [1]. In 2024, an estimated 10.7 million people developed TB, and 1.2 million died [1]. Although TB incidence and mortality have decreased since 2015, progress is far slower than global targets [2]. Each year, about 40% of TB cases remain undetected or unreported [1].

Cambodia, a high-TB burden Southeast Asian nation, faces similar challenges. More than one-third of people with TB have been missed, reflecting major gaps in access to care and accurate diagnosis [1, 3]. TB services are mainly available through public facilities, which are not typically the preferred initial healthcare choice [4]. Another barrier is that >50% of people with TB in high burden settings may not experience typical TB symptoms that would prompt them to seek healthcare [4, 5].

The Community Mobilisation Initiatives to End TB (COMMIT) project was established as a five-year effort to expand access to high-quality, person-centred TB services, improve the TB care delivery system, reduce TB transmission and disease progression, and improve programme outcomes through research and innovation in TB [6]. COMMIT activities included: i) community-based active case finding (ACF) using symptom screening tools and/or chest radiography, ii) TB prevention through TB preventive treatment (TPT) among household contacts of people with bacteriologically confirmed pulmonary TB, and iii) training and capacity building of healthcare workers and peer-support groups.

Although community-based TB interventions are a central pillar of Cambodia’s national strategic plan [7], evidence on their performance in real-world conditions remains limited. The absence of implementation research constrains the ability to integrate high-value practices and de-implement activities with limited benefit. At the same time, the understanding of the epidemic has evolved considerably since COMMIT began, raising concerns about whether current strategies remain well aligned with contemporary needs [8–11].

This gap underscores the need for a systematic assessment of COMMIT’s implementation to determine what worked, what did not, and how programmes could adapt in the future. Using an implementation science framework, this study evaluated the implementation impact of COMMIT, key bottlenecks and challenges, and offered actionable recommendations for future programmes and policies.

## Methods

### Setting

Between 1 October 2019 and 30 September 2024, COMMIT was implemented in four provinces (Phnom Penh, Kandal, Tboung Khmum, and Kampong Cham) and across 27 operational districts (**Figure 1).** COMMIT was implemented by the Khmer HIV/AIDS NGO Alliance (KHANA) in collaboration with the National Centre for Tuberculosis and Leprosy Control and other local non-governmental organisations, namely the Cambodia Anti-Tuberculosis Association (CATA), Health and Social Development (HSD), and Cambodian Health Committee (CHC).

**Figure 1:**
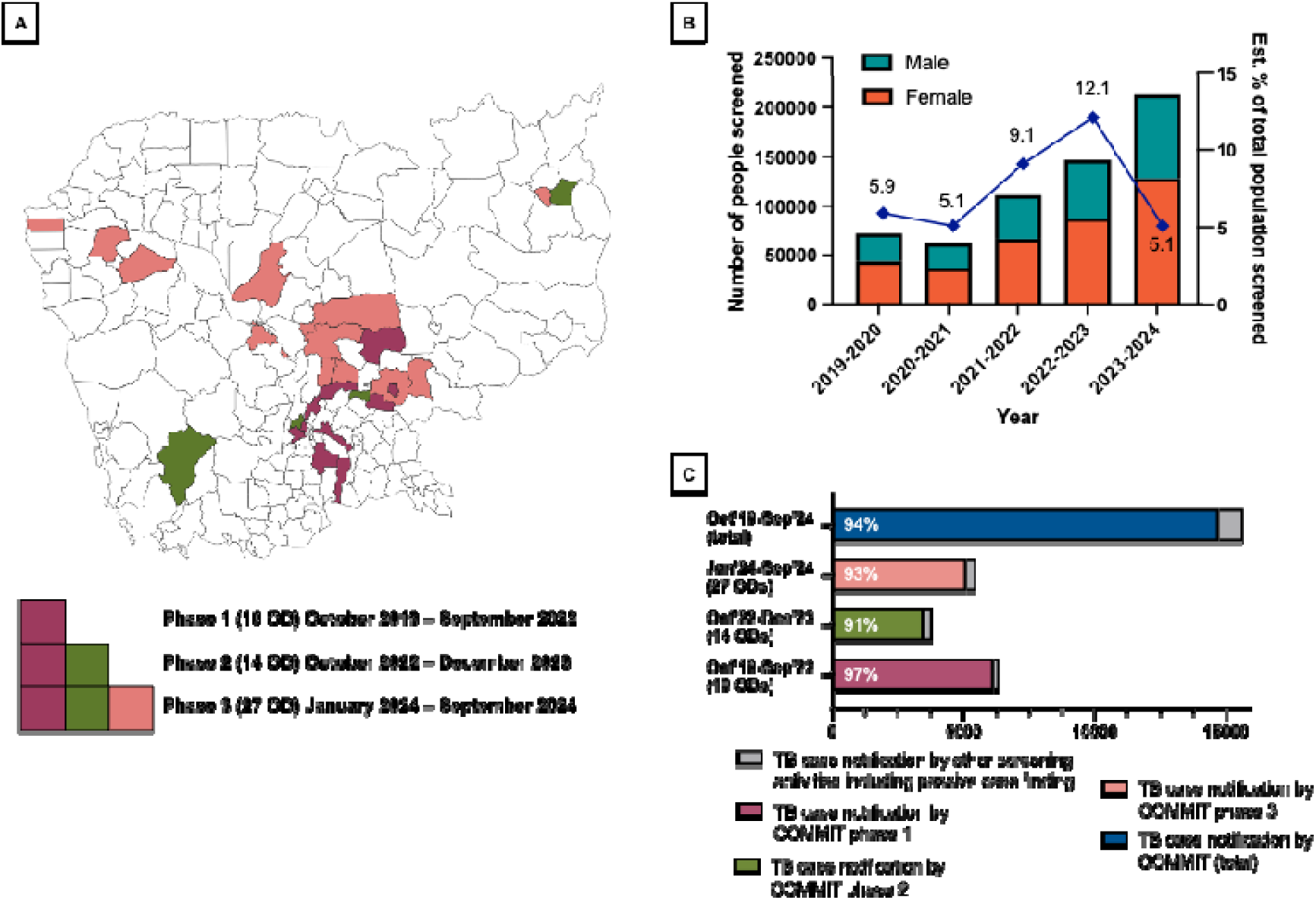
COMMIT coverage and reach. **[A] Map of COMMIT sites at the operational districts level.** Phase 1 included 10 ODs: Kang Meas, Leuk Dek, Lvea Em, Muk Kampoul, Ou Raing Euv, Por Sen Chey, Sa Ang, Sen Sok, Stung Trang, Suong. Phase 2 included 4 additional ODs (total 14 ODs): Bar Kaev, Koh Soutin, Prek Pnov, Srae Ambel. Phase 3 included an additional 13 OD (total 27 ODs): Banlung, Baray-Santuk, Boribo, Cheung Prey, Dambae, Kampong Chhnang, Kampong Thom, Prey Chhor, Sampov Loun, Sangkae, Staung, Thboung Khmum, and Thmar Kol. **[B] Number of people screened for tuberculosis by sex and the estimated proportion of the total population screened**. Year 1 (2019-2020): 10 operational districts, estimated total population = 1.2 million; Year 2 (2020-2021): 10 operational districts, estimated total population = 1.2 million; Year 3 (2021-2022): 10 operational districts, estimated total population = 1.2 million; Year 4 (2022-2023): 14 operational districts, estimated total population = 1.5 million; Year 5 (2023-2024): 27 operational districts, estimated total population = 4.2 million. **[C] Notifications in the national tuberculosis management information system.** The percentages represent the proportion of case notifications contributed by COMMIT to the national TB surveillance system in the respective phases.

### COMMIT programme

COMMIT was designed to target populations at high risk of TB (e.g., TB close contacts, people living with HIV, people with diabetes) in under-resourced communities in Cambodia. The programme implemented four key TB screening models, comprising:

**i.** *<u>Seed-and-recruit model</u>:* Lay counsellors identified persons newly diagnosed with TB and TB survivors in the community as “seeds.” The seeds were trained to recruit individuals with TB symptoms from their social networks. Seeds were provided with basic TB knowledge, communication skills, and referral procedures. The referred individuals underwent diagnostic testing at public health centres or hospitals in accordance with national guidelines [12].
**ii.** *<u>Mobile chest X-ray model</u>:* Mobile diagnostic units equipped with digital chest X-ray and staffed by trained personnel conducted one-off screening events in selected communities. Participants were screened for TB symptoms and underwent chest radiography on-site. Images were interpreted immediately, and sputum samples from individuals with abnormal radiographic findings were collected for on-site GeneXpert testing [12].
**iii.** *<u>Community DOTS (CDOTS):</u>* While the traditional Directly Observed Treatment Short-course (DOTS) focuses on supervising medication for diagnosed patients, CDOTS functioned as an outreach model centred on community-based TB screening. Village Health Support Groups (VHSGs) were engaged and trained to conduct door-to-door screening for TB symptoms within their communities. VHSGs provided TB education, identified people with presumptive TB, and referred them to health centres for diagnostic testing [13].
**iv.** *<u>Hospital linkage model</u>:* Referral hospitals implemented systematic TB symptom screening for all patients attending inpatient wards and outpatient clinics. Health workers used standardised symptom checklists to identify people with presumptive TB. Those identified were referred for diagnostic testing within the hospital or at affiliated TB diagnostic centres. Screening was integrated into routine hospital workflows to maximise coverage [14].

### Study design and evaluation frameworks

We used a mixed-methods explanatory sequential study design [15] to evaluate the implementation of the COMMIT programme. This approach involved a retrospective review of data collected from the programme and the national TB Management Information System (TB-MIS), followed by in-depth interviews and focus group discussions with key stakeholders. This evaluation was structured using the RE-AIM framework to assess implementation efforts at the individual (Reach and Effectiveness) and organisational (Adoption, Implementation, and Maintenance) levels [16–18].

#### Reach

We assessed programme reach by documenting the number of villages, operational districts, and provinces where activities were implemented. Reach was also evaluated by estimating screening coverage among the total population in COMMIT operational districts. We qualitatively explored the rationale behind the observed geographical and population reach, including reasons for limited coverage where applicable.

#### Effectiveness

Effectiveness was evaluated across three key domains: screening, testing, and prevention. For TB screening and testing, we reported the number of individuals screened for TB, those identified as presumptive TB cases (presented with either (1) ³1 World Health Organisation (WHO) 4-symptom screen: fever, weight loss, cough, and night sweats; or 2) an abnormal chest X-ray suggestive of TB, and those tested using either rapid molecular diagnostics or smear microscopy. We also documented the number of individuals diagnosed with TB (all forms) and those successfully treated (cured or treatment completed), according to WHO definitions. Results were disaggregated by model: seed-and-recruit, mobile chest X-ray, CDOTS, and hospital linkage. We also described the number of TB cases identified by COMMIT and notified to the National TB Programme via TB-MIS. It is noteworthy that no other TB interventions were implemented in these sites during the study period.

For TB prevention, we described the number of people eligible for TPT, defined as 1) at high risk of developing active TB and 2) with no active TB as determined by the WHO 4-symptom screen. Next, we documented the number of eligible individuals who enrolled in and initiated TPT, defined as those who received TPT using either 3HP (Weekly Isoniazid and Rifapentine for three months), 3RH (Daily Rifampicin and Isoniazid for three months), or 6H (Daily Isoniazid for six months). We also reported the number of individuals who completed all required doses in accordance with national guidelines.

#### Adoption

We assessed adoption at the setting level by recording the number of villages and health centres invited and that ultimately joined the programme. Staff-level adoption was assessed by describing the number of trained healthcare workers and the number of peer support groups established. We also explored the reasons for adoption or non-adoption.

#### Implementation

Implementation was assessed across the domains of screening, testing, and treatment initiation, and contact investigation. First, we compared screening algorithms across screening models and described variation in symptom-based screening. Implementation was next assessed by describing the proportion of people with presumptive TB, including those with chest X-ray abnormalities, who received a confirmatory TB test. Notable changes in confirmatory diagnostics during implementation were also described. In addition, we calculated the proportion of individuals diagnosed with TB who successfully initiated treatment. We also documented the total number of people with bacteriologically confirmed TB and determined the proportion of household contacts who underwent contact investigation. Key implementation successes and challenges encountered during programme delivery were explored qualitatively.

#### Maintenance

Maintenance was primarily assessed using qualitative methods. We examined programme successes, highlighting key innovations that were routinised into practice and their perceived impacts on national TB elimination goals. We also considered low-value practices for de-implementation [19] and discussed recommendations to sustain or scale up the programme, as well as opportunities for improvement.

### Participant sampling and recruitment

The key stakeholders in this programme included COMMIT implementation partners (KHANA, HSD, CHC, CATA), policymakers (National TB Programme and the local provincial authorities), field TB officers and supervisors, healthcare workers (referral hospitals and health centres), and members of the affected communities in the project sites. These stakeholder groups were pre-identified during the multisectoral programme design stage and were directly involved in programme implementation, monitoring, and evaluation [20]. Participants in the qualitative study were purposively selected through maximum variation sampling from these key stakeholder groups. We included individuals aged 18 or older who had participated in the COMMIT programme for at least three months. To capture a broad spectrum of perspectives and ensure comprehensive coverage, we aimed to recruit five to ten individuals per group, representing diversity in age, sex, experience, affiliations, and geographic locations aligned with the project sites [21].

### Data collection

We collected quantitative data on the key performance indicators and treatment care cascade from the programme database and the national TB-MIS. To complement the quantitative analyses, we applied constructs from the Theoretical Domains Framework (TDF) [22] to explore barriers and enablers to adoption and adherence to planned activities, and to identify factors that might explain variations in effectiveness. Findings from the quantitative component informed the selection of specific observations for further investigation using TDF constructs [15]. We conducted in-depth, semi-structured interviews and focus group discussions to explore the various components of the RE-AIM framework. The interview guide was developed based on preliminary quantitative findings and relevant TDF domains and was pilot-tested before implementation. Interviews were conducted with key stakeholders to gain insights into contextual and practical factors influencing programme implementation, including successes and challenges. We also interviewed members of the affected communities to better understand their experiences, satisfaction with the programme, and suggestions for improvement. All interviews were conducted in the interviewees’ preferred language and audio**-**recorded for analysis.

### Data analyses

We described categorical data using frequencies and percentages. We disaggregated the data by age and sex, as appropriate. To estimate screening coverage, we calculated the proportion of residents who registered for TB screening (across all screening models) by dividing the number of registrants by the estimated population across all ODs [23]. This analysis was further stratified by sex. We also constructed TB and TPT care cascades. The TB care cascade was constructed by calculating the proportion of individuals retained at each step compared with the preceding step. Diagnostic yield was calculated as the number of TB cases identified divided by the number of individuals screened. The TPT cascade was constructed by calculating the proportion of individuals retained at each step relative to the first step (i.e., the number of people eligible for TPT).

Interview transcripts were transcribed and translated into English for analysis. Translated transcripts were reviewed against the original version. Four independent analysts (BS, MN, HT, AKJT) retrieved texts on the topics of interest and categorised them using thematic analysis. We based the initial constructs on the overarching frameworks and developed a codebook accordingly. Any emerging codes were added to the codebook. Coders met to develop the codebook, discuss disagreements, and reach consensus on coding procedures and constructs. Upon completion of coding, qualitative results, including exemplar quotes and quantitative data, were consolidated into a final matrix structured around the RE-AIM and TDF components. We drew conclusions through the triangulation of quantitative and qualitative findings.

#### Ethics

The National Ethics Committee for Health Research in Cambodia (NECHR) (reference: 430/NECHR) approved the study. A waiver of consent was approved for the secondary analysis of existing databases. Written informed consent was obtained from all interview participants.

## Results

### Characteristics of study participants

Demographic characteristics of qualitative respondents are summarised in Table 1. In total, 27 IDIs and two FGDs (with 14 participants) were conducted. Approximately 70% of the participants were male. Participants were distributed across Phnom Penh (63.4%) and four provinces (36.6%).

**Table 1:** Demographic characteristics of qualitative respondents.

|  | n (%) |
| --- | --- |
| Sex |  |
| Male | 29 (70.7) |
| Female | 12 (29.3) |
| Location |  |
| Phnom Penh | 26 (63.4) |
| Kandal | 9 (22.0) |
| Tboung Khmum | 3 (7.3) |
| Kampong Thom | 2 (4.9) |
| Ratanakiri | 1 (2.4) |
| Stakeholder Type |  |
| NGO project staff | 9 (22.0) |
| HC/RH TB officers | 6 (14.6) |
| OD TB supervisors | 4 (9.8) |
| Healthcare workers | 4 (9.8) |
| Policymakers | 3 (7.3) |
| Members of affected communities* | 14 (34.1) |
| Other | 1 (2.4) |

\*Two FGDs were conducted with members of affected communities, each comprised of 6-8 participants. Abbreviations: HC (health centre), OD (operational district), RH (referral hospital), TB (tuberculosis)

### Reach

COMMIT covered nine of the 25 provinces (36%), 27 of the 103 operational districts (26.2%), and 1,509 of the 14,577 villages (10.4%) and screened approximately 5-12% of the population in participating locations. A higher proportion of women (60%) than men (40%) attended the screening activities.

The geographical coverage and site selection were determined by funding and operational constraints within the National TB Programme, particularly in areas without ongoing active case-finding interventions. The programme’s reach focused on key populations prioritised by the national programme.

> *“The target groups we are working with include all key populations identified in the national programme, such as those at risk and affected by TB, diabetes, individuals over 55 years old, malnourished children, migrant workers, people living in urban slums, and people in prison. Specifically, we concentrate on individuals who live closely with those who have TB.” (IDI05, NGO)*

Reaching target populations was limited by poor knowledge and negative beliefs about TB, particularly among older adults and men. Many older adults felt physically unable to participate in screening, and men tended to neglect symptoms and delay care. Participation was also reduced by daytime scheduling of screening activities and stigma, particularly fears of income loss or damaged reputation. Hard-to-reach groups, including people in prisons and factory and construction workers, remained underserved due to logistical and structural barriers (**Table 2**).

**Table 2:**
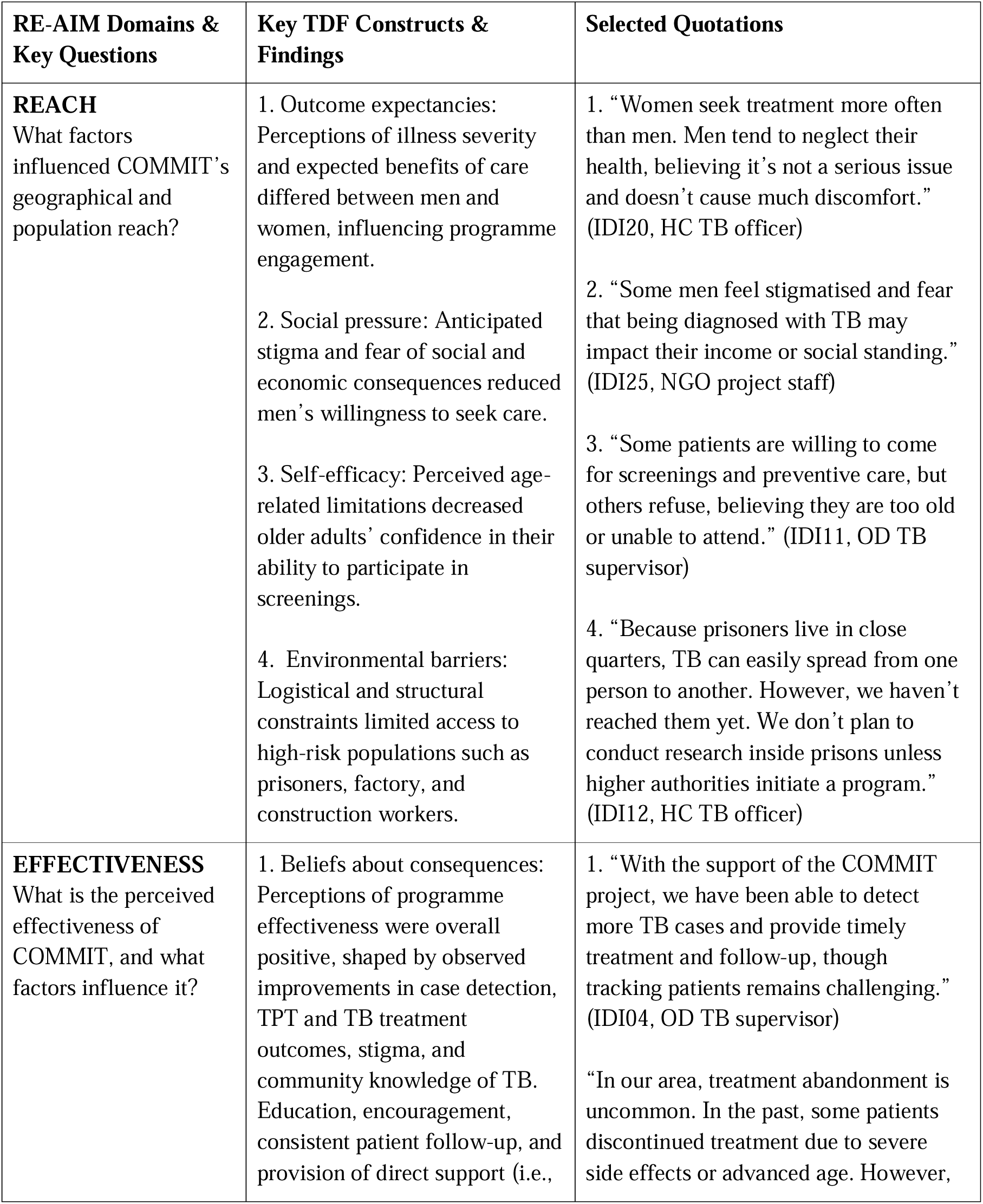
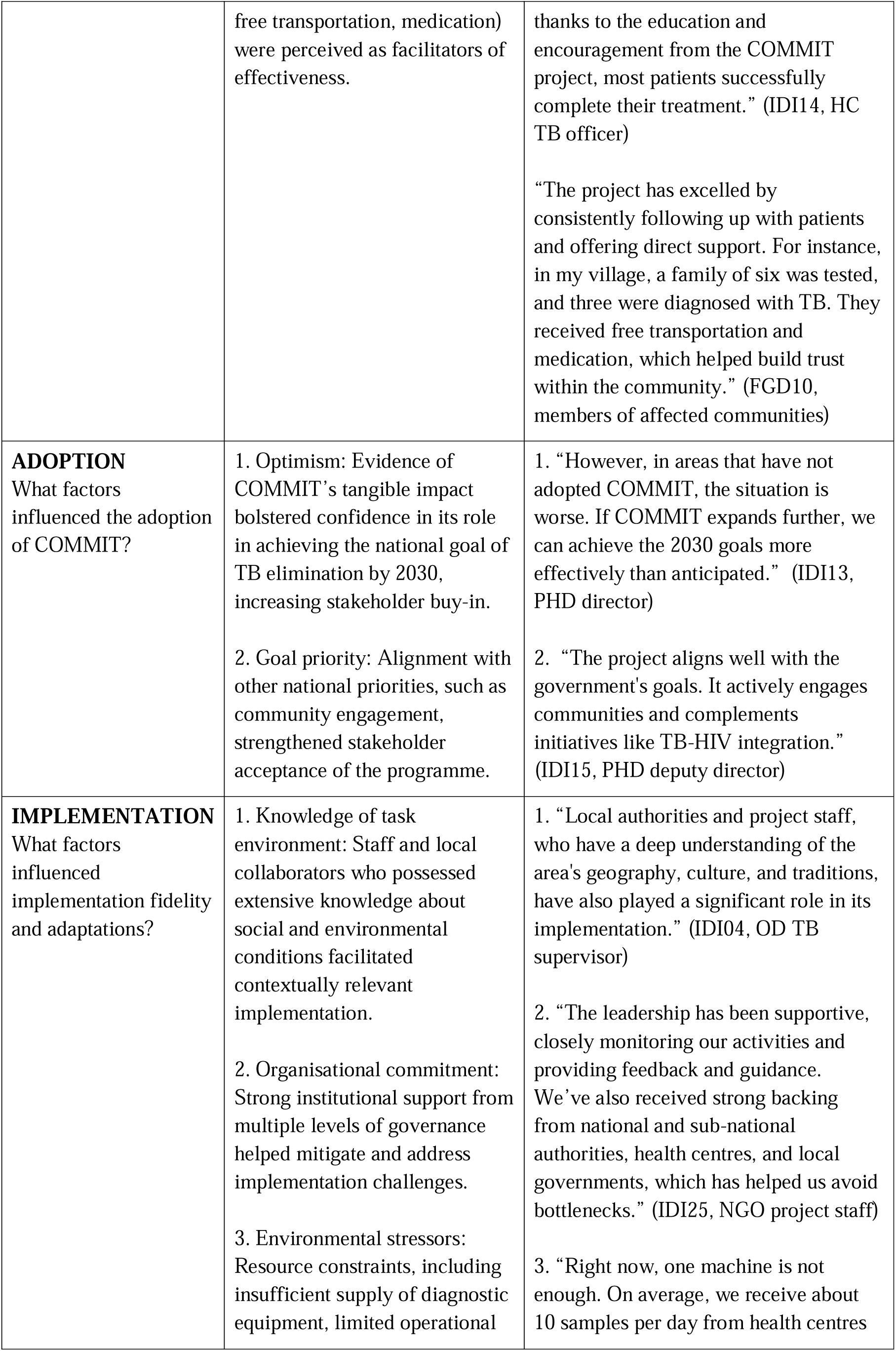
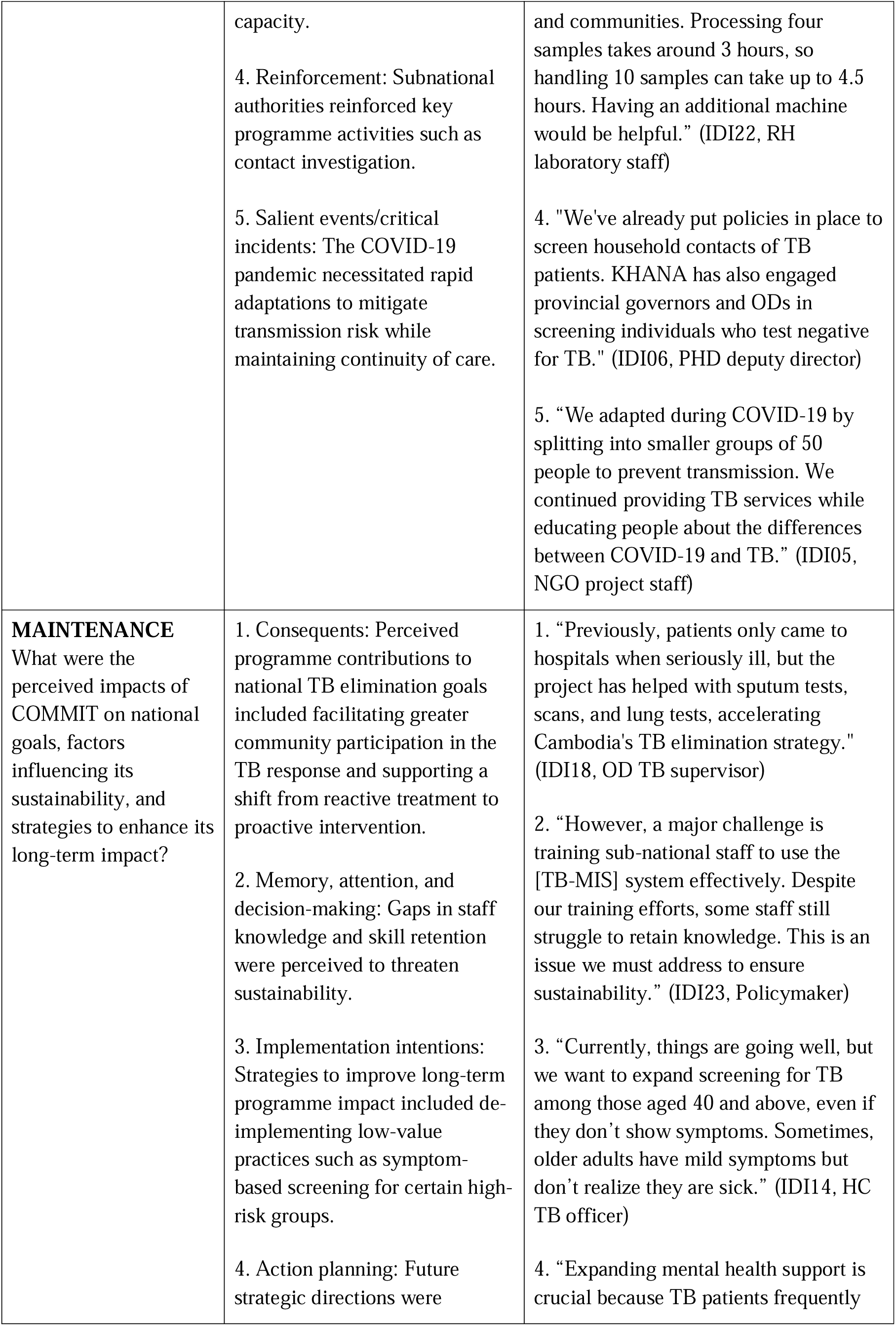
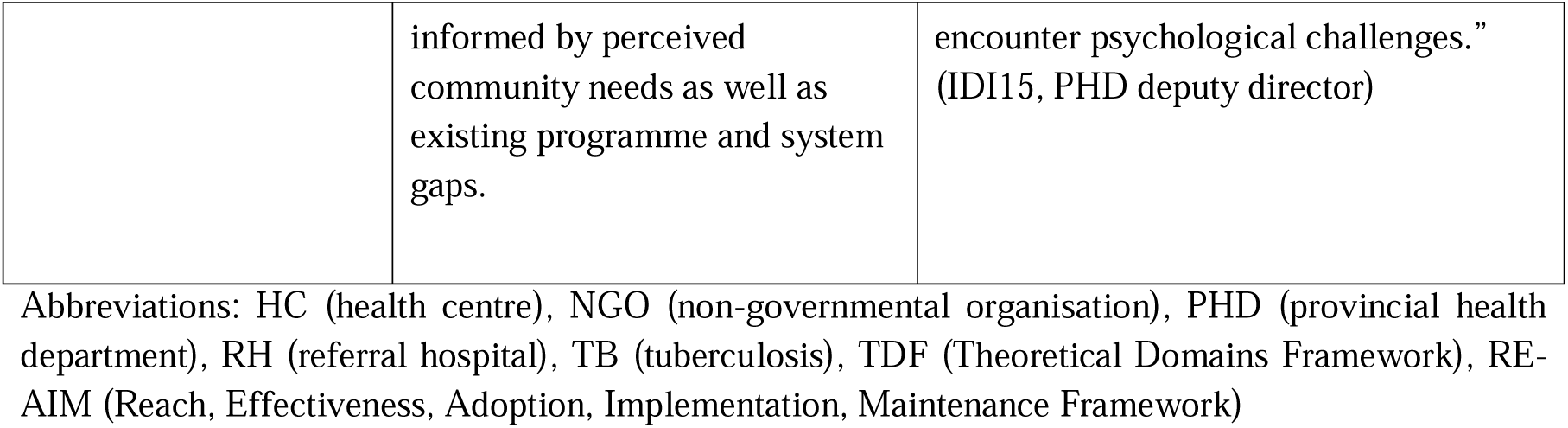
Selected qualitative quotes for key TDF constructs and RE-AIM framework.

| RE-AIM Domains & Key Questions | Key TDF Constructs & Findings | Selected Quotations |
| --- | --- | --- |
| **REACH**<br>What factors influenced COMMIT's geographical and population reach? | 1. Outcome expectancies: Perceptions of illness severity and expected benefits of care differed between men and women, influencing programme engagement.<br>2. Social pressure: Anticipated stigma and fear of social and economic consequences reduced men's willingness to seek care.<br>3. Self-efficacy: Perceived age-related limitations decreased older adults' confidence in their ability to participate in screenings.<br>4. Environmental barriers: Logistical and structural constraints limited access to high-risk populations such as prisoners, factory, and construction workers. | 1. "Women seek treatment more often than men. Men tend to neglect their health, believing it's not a serious issue and doesn't cause much discomfort." (IDI20, HC TB officer)<br>2. "Some men feel stigmatised and fear that being diagnosed with TB may impact their income or social standing." (IDI25, NGO project staff)<br>3. "Some patients are willing to come for screenings and preventive care, but others refuse, believing they are too old or unable to attend." (IDI11, OD TB supervisor)<br>4. "Because prisoners live in close quarters, TB can easily spread from one person to another. However, we haven't reached them yet. We don't plan to conduct research inside prisons unless higher authorities initiate a program." (IDI12, HC TB officer) |
| **EFFECTIVENESS**<br>What is the perceived effectiveness of COMMIT, and what factors influence it? | 1. Beliefs about consequences: Perceptions of programme effectiveness were overall positive, shaped by observed improvements in case detection, TPT and TB treatment outcomes, stigma, and community knowledge of TB. Education, encouragement, consistent patient follow-up, and provision of direct support (i.e., | 1. "With the support of the COMMIT project, we have been able to detect more TB cases and provide timely treatment and follow-up, though tracking patients remains challenging." (IDI04, OD TB supervisor)<br>"In our area, treatment abandonment is uncommon. In the past, some patients discontinued treatment due to severe side effects or advanced age. However, |
|  | free transportation, medication) were perceived as facilitators of effectiveness. | thanks to the education and encouragement from the COMMIT project, most patients successfully complete their treatment.” (IDI14, HC TB officer)<br>“The project has excelled by consistently following up with patients and offering direct support. For instance, in my village, a family of six was tested, and three were diagnosed with TB. They received free transportation and medication, which helped build trust within the community.” (FGD10, members of affected communities) |
| **ADOPTION**<br>What factors influenced the adoption of COMMIT? | 1. Optimism: Evidence of COMMIT’s tangible impact bolstered confidence in its role in achieving the national goal of TB elimination by 2030, increasing stakeholder buy-in.<br>2. Goal priority: Alignment with other national priorities, such as community engagement, strengthened stakeholder acceptance of the programme. | 1. “However, in areas that have not adopted COMMIT, the situation is worse. If COMMIT expands further, we can achieve the 2030 goals more effectively than anticipated.” (IDI13, PHD director)<br>2. “The project aligns well with the government's goals. It actively engages communities and complements initiatives like TB-HIV integration.” (IDI15, PHD deputy director) |
| **IMPLEMENTATION**<br>What factors influenced implementation fidelity and adaptations? | 1. Knowledge of task environment: Staff and local collaborators who possessed extensive knowledge about social and environmental conditions facilitated contextually relevant implementation.<br>2. Organisational commitment: Strong institutional support from multiple levels of governance helped mitigate and address implementation challenges.<br>3. Environmental stressors: Resource constraints, including insufficient supply of diagnostic equipment, limited operational | 1. “Local authorities and project staff, who have a deep understanding of the area's geography, culture, and traditions, have also played a significant role in its implementation.” (IDI04, OD TB supervisor)<br>2. “The leadership has been supportive, closely monitoring our activities and providing feedback and guidance. We’ve also received strong backing from national and sub-national authorities, health centres, and local governments, which has helped us avoid bottlenecks.” (IDI25, NGO project staff)<br>3. “Right now, one machine is not enough. On average, we receive about 10 samples per day from health centres |
|  | capacity.<br>4. Reinforcement: Subnational authorities reinforced key programme activities such as contact investigation.<br>5. Salient events/critical incidents: The COVID-19 pandemic necessitated rapid adaptations to mitigate transmission risk while maintaining continuity of care. | and communities. Processing four samples takes around 3 hours, so handling 10 samples can take up to 4.5 hours. Having an additional machine would be helpful.” (IDI22, RH laboratory staff)<br>4. "We've already put policies in place to screen household contacts of TB patients. KHANA has also engaged provincial governors and ODs in screening individuals who test negative for TB." (IDI06, PHD deputy director)<br>5. “We adapted during COVID-19 by splitting into smaller groups of 50 people to prevent transmission. We continued providing TB services while educating people about the differences between COVID-19 and TB.” (IDI05, NGO project staff) |
| **MAINTENANCE**<br>What were the perceived impacts of COMMIT on national goals, factors influencing its sustainability, and strategies to enhance its long-term impact? | 1. Consequents: Perceived programme contributions to national TB elimination goals included facilitating greater community participation in the TB response and supporting a shift from reactive treatment to proactive intervention.<br>2. Memory, attention, and decision-making: Gaps in staff knowledge and skill retention were perceived to threaten sustainability.<br>3. Implementation intentions: Strategies to improve long-term programme impact included de-implementing low-value practices such as symptom-based screening for certain high-risk groups.<br>4. Action planning: Future strategic directions were | 1. “Previously, patients only came to hospitals when seriously ill, but the project has helped with sputum tests, scans, and lung tests, accelerating Cambodia's TB elimination strategy.” (IDI18, OD TB supervisor)<br>2. “However, a major challenge is training sub-national staff to use the [TB-MIS] system effectively. Despite our training efforts, some staff still struggle to retain knowledge. This is an issue we must address to ensure sustainability.” (IDI23, Policymaker)<br>3. “Currently, things are going well, but we want to expand screening for TB among those aged 40 and above, even if they don’t show symptoms. Sometimes, older adults have mild symptoms but don’t realize they are sick.” (IDI14, HC TB officer)<br>4. “Expanding mental health support is crucial because TB patients frequently |
|  | informed by perceived community needs as well as existing programme and system gaps. | encounter psychological challenges.” (IDI15, PHD deputy director) |

Abbreviations: HC (health centre), NGO (non-governmental organisation), PHD (provincial health department), RH (referral hospital), TB (tuberculosis), TDF (Theoretical Domains Framework), RE-AIM (Reach, Effectiveness, Adoption, Implementation, Maintenance Framework)

### Effectiveness

#### TB care cascade

Overall, more than 690,000 individuals were screened for TB using either symptom assessment or chest X-ray as the initial screening tool. Among them, 25% screened positive, and approximately 98% underwent diagnostic testing with GeneXpert or smear microscopy. Of those tested, 10.3% were diagnosed with TB. Overall, 2.5% of individuals screened for TB were diagnosed with TB (all forms), and more than 90% were successfully treated, achieving either treatment completion or cure **(Figure 2A).**

**Figure 2:**
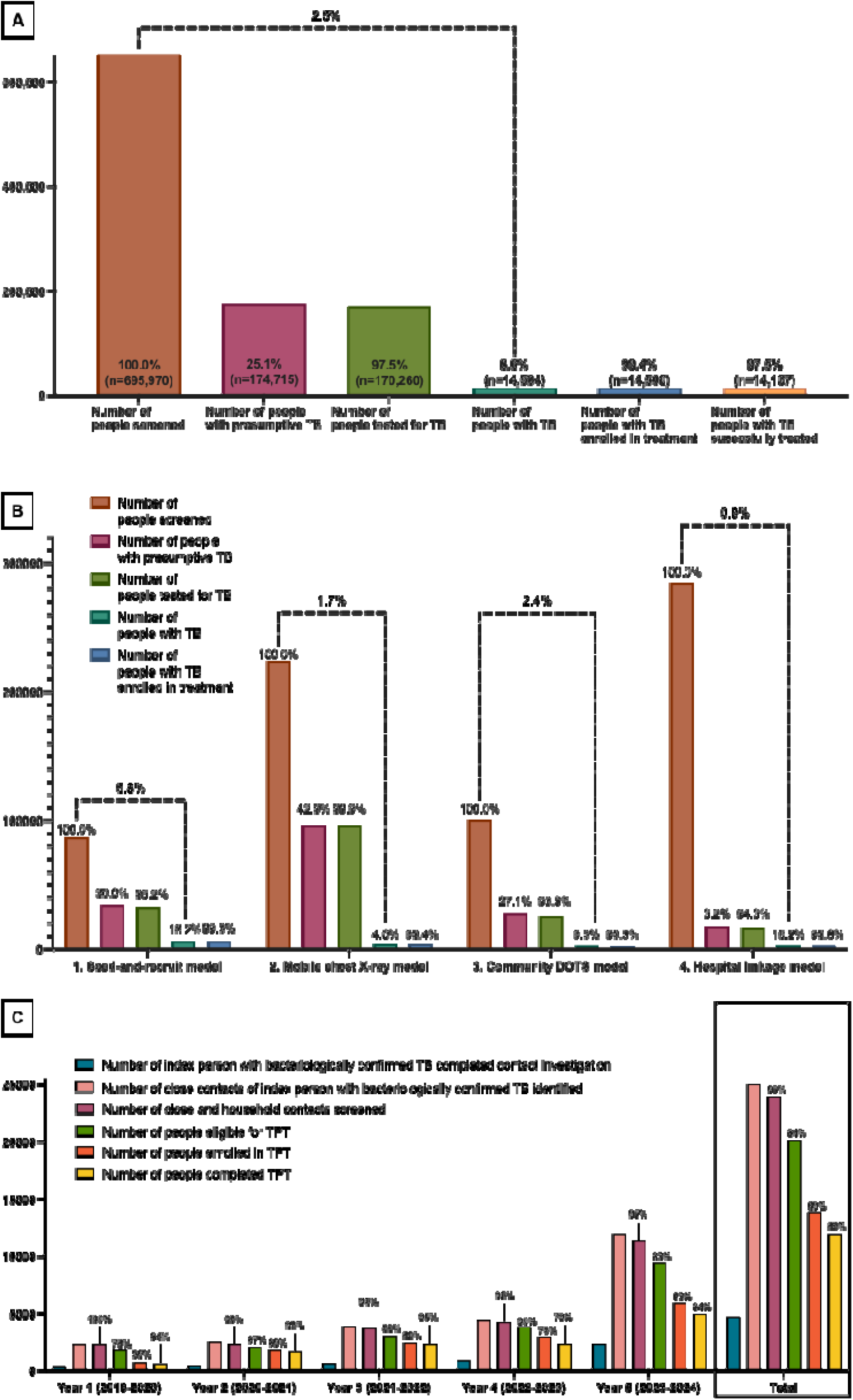
COMMIT TB care cascade. **[A] Care cascade for the total number of people screened by the COMMIT programme. [B] Tuberculosis disease care cascade by screening models.** The percentages were calculated using the denominators from the preceding step. The dotted lines represent the proportion of screened individuals who were confirmed to have TB (all forms). **[C] TB preventive treatment care cascade.** The percentages were calculated using the denominators from the preceding step, except for the first two steps. The number of index persons with bacteriologically confirmed TB who completed contact investigation was 407 (year 1), 469 (year 2), 622 (year 3), 873 (year 4), 2342 (year 5), 4713 (total). The number of close contacts (including household contacts) of index persons with bacteriologically confirmed TB identified was 2316 (year 1), 2515 (year 2), 3878 (year 3), 4401 (year 4), 11930 (year 5), 25040 (total). Abbreviations: COMMIT (Community Mobilisation Initiatives to End TB), TB (tuberculosis), TPT (tuberculosis preventive treatment).

The four screening models differed in screening coverage and diagnostic yield. The hospital linkage model had the highest screening coverage (284,686), followed by the mobile chest X-ray model (223,874), the community DOTS model (100,505), and the seed-and-recruit model (86,905). Conversely, the hospital linkage model had the lowest diagnostic yield (0.9%) compared to the mobile chest X-ray (1.7%), CDOTS (2.4%), and seed-and-recruit (6.4%) models **(Figure 2B)**.

Across the four models, 14,718 cases were detected. Cases detected through COMMIT accounted for 94.1% of all case notifications in the ODs covered by the programme **(Figure 1C)**. Over the duration of the project, COMMIT identified 10% of the nationwide cumulative total of tuberculosis cases.

#### TPT care cascade

Over the project period, 20,104 individuals were identified as eligible for TPT. Of those eligible, 13,814 (68.7%) enrolled in TPT, and 11,912 (86.2%) completed the regimen **(Figure 2C)**.

### Adoption

All invited villages and health centres within the selected ODs participated in the COMMIT programme. Qualitative participants attributed the adoption to its alignment with national priorities, particularly its emphasis on community engagement. Participants also noted that ODs implementing COMMIT demonstrated higher uptake of TB services than non-adopting areas. Its demonstrated impact increased confidence in the programme and strengthened stakeholder support for its role in the 2030 end-TB goal **(Table 2)**.

> *“The results indicated that we have effectively utilised community engagement and made a direct impact on those affected by TB. We actively seek out people with TB, provide care, and ensure access to treatment. The United States Agency for International Development (USAID) has enabled us to expand our target areas. In the first three years, we operated in 10 ODs. By the fourth year, we expanded to 14 ODs, and in the fifth year, we added 13 more, bringing the total to 27 ODs.” (IDI05, NGO COMMIT project staff)*

The programme’s expansion was supported by staff-level adoption from healthcare workers and peer support group members. The number of trained healthcare workers increased significantly over the project period, from 274 in 2019–2020 to 1,923 in 2023–2024. The number of peer support groups and their members also rose substantially, reaching 150 groups and 2,770 members by the final year (**Table 3**).

**Table 3:** Number of healthcare workers who received training and the number of peer support groups established.

|  | 2019-2020 | 2020-2021 | 2021-2022 | 2022-2023 | 2023-2024 |
| --- | --- | --- | --- | --- | --- |
| **Number of healthcare workers who received training (cumulative)** |  |  |  |  |  |
| Female | 90 | 420 | 841 | 944 | 1382 |
| Male | 184 | 902 | 1210 | 1257 | 1306 |
| Total | 274 | 1322 | 2051 | 2201 | 2688 |
| **Peer support group establishment** |  |  |  |  |  |
| Number of groups | 2 | 56 | 73 | 100 | 150 |
| Number of members | 16 | 725 | 1091 | 1556 | 2770 |

### Implementation

#### Screening algorithms

Screening processes varied across ACF models, including symptom-based screening. The seed-and-recruit, CDOTS, and hospital linkage models used symptom-based screening for all age groups and subsequently conducted chest X-rays for symptomatic individuals. In contrast, the mobile ACF model screened all individuals aged 55 and older by chest X-ray, regardless of symptoms. Among those screened under this symptom-independent approach, 13.5% (443/3282) of confirmed cases were asymptomatic **(Table 4)**.

**Table 4:** Characteristics of cases aged ≥55 detected via the mobile active case-finding model.

|  | **Bacteriologically-confirmed TB** | **Clinically-diagnosed TB** | **Extrapulmonary TB** | **Total** |
| --- | --- | --- | --- | --- |
|  | **n (%)** | **n (%)** | **n (%)** | **n (%)** |
| **Age** |  |  |  |  |
| 55 – 64 | 370 (30.5) | 482 (23.3) | 0 (0.0) | 852 (26.0) |
| $\geq 65$ | 843 (69.5) | 1586 (76.7) | 1 (100.0) | 2430 (74.0) |
| **Sex** |  |  |  |  |
| Male | 723 (59.6) | 967 (46.8) | 0 (0.0) | 1690 (51.5) |
| Female | 490 (40.4) | 1101 (53.2) | 1 (100.0) | 1592 (48.5) |
| **Chest X-rays** |  |  |  |  |
| Normal | 0 (0.0) | 0 (0.0) | 1 (100.0) | 1 (0.0) |
| Abnormal not TB | 100 (8.2) | 0 (0.0) | 0 (0.0) | 100 (3.0) |
| Abnormal TB | 1113 (91.8) | 2068 (100.0) | 0 (0.0) | 3181 (96.9) |
| **Symptoms\* reported** |  |  |  |  |
| Yes | 1054 (86.9) | 1784 (86.3) | 1 (100.0) | 2839 (86.5) |
| No | 159 (13.1) | 284 (13.7) | 0 (0.0) | 443 (13.5) |
| **Total** | 1213 | 2068 | 1 | 3282 |

Abbreviation: TB (tuberculosis).
\*Based on the WHO TB 4-symptom screen (cough, fever, night sweats, and weight loss)

#### Confirmatory testing and treatment initiation

A shift in diagnostic methods over the project period was observed, driven by the implementation and scale-up of rapid molecular diagnostics. The programme began with 10 Xpert machines in 2020 and increased to 27 by 2024, achieving coverage across all COMMIT ODs. As capacity increased, Xpert testing replaced smear microscopy as the primary modality for bacteriological confirmation, with smear microscopy used mainly for treatment follow-up. Xpert tests accounted for 96.4% (143,244) of the 147,626 total confirmatory tests conducted over the project period, whereas smear microscopy accounted for only 4,382 (3.6%).

Overall, 97.5% (170,260/174,715) of people with presumptive TB were tested for TB. Rates of confirmatory testing of presumptive TB exceeded 90% in all four ACF models, ranging from 93.3% in the CDOTS model to 99.9% in the Mobile ACF model. In addition, treatment initiation rates were approximately 99% in all models **(Figure 2B)**.

These patterns were supported by streamlined sample referral procedures, rapid reporting of results, and prompt linkage to treatment following diagnosis. Grassroots delivery strategies, such as decentralised linkage to care, supported by a strong cadre of community-based staff and volunteers, reinforced adherence to the diagnostic and treatment pathway.

> *“When a case is suspected, the samples are sent to our laboratory for GeneXpert testing. If the result is positive, we promptly notify the relevant health centres, referral hospitals, and organisations. We inform them of the positive result and request that they initiate treatment immediately to reduce transmission to others.” (IDI22, referral hospital laboratory staff)*

> *“Strong collaboration and effective community volunteer selection have allowed us to clearly identify people with TB and provide successful treatment within their communities. (IDI13, provincial health department director)*

However, high testing uptake revealed bottlenecks in diagnostic capacity. Following the scale-up of Xpert to one machine per OD, certain sites reported that reliance on a single machine was insufficient for meeting growing testing demand **(Table 2)**.

#### Contact investigation

Contact investigation was completed for 4,713 TB index cases. Overall, 25,040 close contacts were notified, including 18,764 household members. More than 93% of notified contacts were screened for TPT eligibility each year **(Figure 2C)**.

Contact investigation efforts were aided by strong backing from local authorities. OD TB supervisors assisted in identifying relevant contacts of TB index cases, and provincial health departments reinforced contact screening efforts **(Table 2)**.

> *“When we find someone with TB, we also screen their family members since TB is contagious. If a family member is infected, they can spread it; therefore, we compile a list of all relevant individuals for prevention and treatment.” (ID04, OD TB supervisor)*

#### Adaptations during the COVID-19 pandemic

COMMIT was implemented during the COVID-19 pandemic, necessitating rapid adaptations to maintain operations while minimising risk. This included physical distancing measures to reduce transmission, provision of personal protective equipment, and transitions to hybrid and remote modalities for patient follow-up and education.

> *“During the COVID-19 pandemic, we reduced direct contact and only conducted screenings for suspected cases. We provided protective equipment and followed up remotely whenever possible.” (IDI11, OD TB supervisor)*

Staff also adapted routine programming to include education on distinguishing between TB and COVID-19, given their overlapping symptoms **(Table 2)**. Additionally, to counter widespread fear and hesitancy about seeking care, staff encouraged people with TB to continue accessing essential health services.

> *“However, many individuals were afraid to visit the health centre due to their concerns about COVID-19. We still encouraged those experiencing symptoms to come in for screening and treatment.” (IDI08, health centre doctor)*

### Maintenance

#### Programme successes

Several TB control interventions were routinised under COMMIT and were perceived to have lasting impacts. This included contact investigation and TPT provision, both of which were scaled up annually **(Figure 2C)**. Participants noted that COMMIT’s emphasis on early case detection and prevention supported a shift from reactive treatment to proactive intervention, thereby supporting progress toward TB elimination goals **(Table 2)**.

> *“Absolutely, it [COMMIT] has a significant impact [on the national goal of TB elimination by 2030]. We recognise that TB is contagious and that it is effectively prevented through patient treatment and by halting new infections. If individuals remain healthy, the disease will decline.” (IDI13, provincial health department director)*

Community mobilisation initiatives, such as community-led monitoring (CLM) [24], were also scaled up over the programme period. CLM capacity expanded as peer support groups and membership increased **(Table 3)**. TB-affected people even assumed responsibilities beyond those of CLM, such as mutual aid organising and spreading TB awareness, reflecting greater community participation in the TB response over time.

> *“Networks of TB patients who have completed their treatment have formed groups to collect donations and assist others in the community. They have even established associations recognised by the Ministry of Interior, becoming active advocates for TB awareness.” (IDI05, NGO project staff)*

#### De-implementation

Qualitative respondents perceived symptom-based screening as unreliable because it relies on patients’ recognition and self-report of symptoms to determine who may proceed to testing.

> *“Sometimes TB bacteria can be dormant and show no symptoms. For example, a person may not cough but might experience a fever at night, thinking it’s normal. They could also eat regularly but still lose weight, which might indicate TB.” (IDI12, health centre TB officer)*

Participants also cautioned against equating the absence of symptoms to the absence of TB disease, given the unpredictable timing of symptom onset and reporting. Due to the subjective nature of symptom-based screening and its potential to miss asymptomatic TB, participants expressed intentions to reduce reliance on this approach to improve case detection. **(Table 2)**.

> *“When the immune system weakens, symptoms may emerge. This means that everyone is at risk of TB; it’s just a matter of when those symptoms will show up.” (IDI07, referral hospital doctor)*

#### Strategic future directions

Strengthening TB information systems to improve surveillance and programme responsiveness emerged as a key recommendation in priority-setting discussions. Participants called for greater investment in human resources to support data management, alongside regular refresher training on TB-MIS to sustain data quality **(Table 2)**. They also advocated for equipping facilities with more technology, phasing out paper-based documentation, and enhancing digital infrastructure to enable real-time case reporting.

> *“In the future, I hope to minimise paperwork and transition to a fully digital system, in line with the government’s digital transformation goals. This will make the process faster and more efficient.” (IDI23, policymaker)*

Moreover, participants underscored the need to enhance private-sector reporting to achieve comprehensive surveillance. Recognising the significant role of private TB services, a participant called for strengthening collaboration between the public and private sectors to reduce fragmentation in TB control efforts.

> *“Public institutions report 100%, while private services often fail to report detected cases. Collaboration between the public and private sectors is essential…We have already engaged in some informal collaboration, but it has not been formalised yet.” (IDI02, NGO project staff)*

Participants also recommended building subnational capacity for financial stewardship, asserting that such decentralisation efforts would help sustain service delivery beyond the programme period.

> *“I recommend enhancing the ability of local authorities to manage resources, particularly the funds allocated for health services. This will help ensure that TB services continue to reach the target groups even after the project concludes.” (IDI26, NGO project staff)*

To maximise future programme impact, participants suggested strengthening bidirectional TB-diabetes screening, citing the benefits of integrated services for treatment outcomes. Participants also proposed expanding integrated services to address other TB comorbidities and risk factors, such as hypertension, mental health, HIV, and undernutrition, based on perceived disease burden and community needs **(Table 2)**.

> *“Screening for both diseases (TB & diabetes) has many benefits. Treating both conditions increases the chances of success. For example, if a patient has both TB and diabetes, treating both leads to better outcomes.” (IDI14, health centre TB officer)*

## Discussion

### Screening, testing, case detection, treatment, and care

Despite reaching multiple priority groups, COMMIT ultimately screened only a small fraction of the general population, reflecting both the programme’s riskDgroupDfocused strategy and the operational constraints that limited geographical coverage. In highDburden contexts such as Cambodia, where TB transmission is likely to be widespread, virtually everyone is at risk [25]. Although targeted screening models may appear highly efficient, yielding a large number of TB cases relative to the number of people screened, they likely represent only the “tip of the iceberg” of the true disease burden. Empirical evidence from Cambodia indicated that TB is not limited to groups with recognised risk factors [26], and very little is known about the characteristics or burden of cases missed through targeted approaches alone. Furthermore, many people with TB in the community may be asymptomatic, making them unlikely to be reached through symptom-gated approaches alone [5, 27].

After more than a decade of targeted ACF efforts, an estimated 30–40% of TB cases in Cambodia continue to be missed each year [1]. These persistent gaps indicate that current predominantly riskDgroup–based strategies may be insufficient. Ultimately, the goal of TB programmes is not merely to detect as many cases as possible within a restricted group, but to identify the people who remain undetected, as these individuals continue to sustain transmission within the community. Therefore, while targeted approaches are operationally efficient, screening strategies will need to expand beyond narrowly defined highDrisk groups to include the general population in high-burden settings [11, 25] to interrupt transmission and reduce the pool of undiagnosed TB.

The uneven screening participation patterns, with lower engagement among men, highlight the influence of limited knowledge, stigma, and structural barriers on healthDseeking behaviour [28, 29]. To expand the programme’s reach, future efforts could strengthen community mobilisation, peer support, and stigma reduction, while decentralising care to improve accessibility for hardDtoDreach populations. Such adaptations are essential to increase participation in screening and ensure that populations most vulnerable to TB are not left behind. These findings emphasise the importance of grounding screening strategies in a nuanced understanding of local epidemiology, including who remains unreached and why.

Despite these limitations and the need to adapt implementation during the COVID-19 pandemic, COMMIT contributed substantially to overall TB case detection, underscoring the overall role of ACF in high-burden settings [30]. Continued investment in ACF using evidence-based approaches will be essential to close persistent detection gaps. The shift from a less sensitive diagnostic tool, such as smear microscopy, to rapid molecular diagnostics during the project period was also crucial to improving case detection. However, the effective implementation of rapid molecular diagnostics depends on a well functioning testing ecosystem, including reliable maintenance, quality assurance, and careful monitoring of supply and demand.

Cambodia has strong treatment indicators, with initiation and success rates consistently above the global average [1]. Where most ACF programmes experienced high pre-treatment attrition rates [31], COMMIT’s community-based peerDsupport model was instrumental in sustaining high rates of treatment initiation and completion. The major programmatic gap lies earlier in the cascade: screening and initiating the care continuum [3]. Ultimately, we cannot control TB unless we find those who have it.

### Contact investigation and TB preventive treatment

Contacts of people with TB are among the groups at highest risk of developing the disease [32]. Current national guidelines, aligned with WHO recommendations, prioritise contact investigation for individuals exposed to bacteriologically confirmed pulmonary TB and drug resistant TB [11, 33]. Despite this, contact investigation has not been systematically implemented across Cambodia [34, 35]. COMMIT filled the gap by supporting contact investigation and evaluation for TPT. In this evaluation, we found that >90% of the notified contacts were screened, and TPT enrolment also far exceeded the national average of 37% [1]. This demonstrated COMMIT’s capacity to operationalise contact investigation and the TPT programme. Integrating this into routine care at scale beyond the COMMIT framework warrants further deliberation.

Nevertheless, relying solely on symptom based screening to determine eligibility for TPT risks missing individuals with asymptomatic TB disease. Recent estimates suggest that fewer than one quarter of the population in Cambodia has been infected with TB [36], consistent with estimates for the wider Western Pacific region [37]. Without routine TB infection testing, many individuals may receive TPT unnecessarily. Together, these challenges suggest that symptomDbased eligibility screening warrants deDimplementation, and that TPT eligibility criteria should be refined and optimised accordingly.

### Local engagement, capacity building, and community mobilisation

Local engagement and capacity building were central to COMMIT’s success and closely aligned with Pillar 2 of the WHO End TB Strategy, which emphasises meaningful participation of communities and civil society in TB response [38]. COMMIT was strongly supported by local authorities, community volunteers, and peer networks to deliver peopleDcentred care. The programme’s expansion from 10 to 27 ODs, and the widespread uptake of activities such as TB screening, contact investigation, and treatment support, demonstrated how a groundDup, locally driven model fosters ownership and improves programme adoption. This approach resonates with global evidence that affected communities are experts in their care pathways, and that engaging them early and substantively enhances the relevance, quality, and sustainability of TB interventions [39, 40].

COMMIT also strengthened community capacity through substantial training for healthcare workers and by expanding peer support groups. These peer networks and valuable resources have evolved beyond COMMIT to other communityDled interventions and TB advocacy at the local and national levels. For instance, they have been instrumental in designing and delivering other major TB interventions in the country [41]. By enabling communities to identify barriers across the TB care cascade and coDcreate solutions with health authorities, COMMIT demonstrated that community participation not only strengthens health system capacity but also creates sustainable mechanisms for TB response beyond the programme’s remit. This model confirms that engaging communities is a critical pathway to advance toward TB elimination.

### Looking forward

We recognise the reality that a programme of this scale requires substantial financial investment. With global health funding declining, Cambodia faces an urgent need to pivot toward more sustainable financing solutions. Currently, only 17% of the national TB budget is domestically funded, compared with 26% from international sources, leaving a funding gap exceeding 50% [1].

Cambodia’s rapidly growing economy and its successes in TB control, supported by significant external funding, make it pivotal for the government to increase domestic investment. Prioritising evidenceDbased interventions and communityDcentred activities would ensure the greatest value for money in the long term. Furthermore, stakeholders consistently reported that COMMIT accelerated Cambodia’s shift from reactive care to proactive case-finding and prevention. Sustaining this impact will require deliberate strategies to preserve learned experience, strengthen decisionDmaking capacities, and ensure continuity of practice among healthcare workers and communityDbased screening and prevention.

The long-term maintenance of the COMMIT model depends on institutionalising its key strengths. This includes strong local ownership and capacity, peerDsupport networks that extend beyond project timelines, and coDcreation of solutions with affected communities to identify and overcome barriers across the care cascade. Future models of care must be personDcentred, responsive to evolving community needs, operationally efficient, and integrated across health conditions. COMMIT focused primarily on TB, but siloed models can overlook the complex realities faced by people with overlapping health challenges. Given the growing synergies between communicable and non-communicable diseases in contexts such as Cambodia, more integrated service delivery approaches can help ensure comprehensive and equitable care [42]. Engagement with other sectors, particularly the private health care sector, which is often the first point of contact for people with TB, remains a key priority of TB response [3]. At the same time, Cambodia should discontinue low-value practices such as exclusive reliance on symptom-gated screening for TB case detection and TPT eligibility determination in the community, given persistent evidence that asymptomatic TB disease is common and often missed with such approaches.

### Limitations

This evaluation has several limitations. The analysis relied on routinely collected programme and TBDMIS data, which may be subject to reporting inconsistencies, variable data quality, limited granularity, and missingness. The study also lacked a control arm, so improvements in case detection and prevention may reflect unmeasured contextual or secular trends rather than the programme alone. Participation in qualitative interviews may have been affected by selection and social desirability biases, as individuals involved in implementing the COMMIT programme may be more inclined to provide favourable views. We attempted to minimise this by using an independent qualitative researcher who was not affiliated with any implementing organisation and who clearly communicated the need for honest, comprehensive feedback. Perspectives from some affected community members may have been underDrepresented, particularly those who were less engaged or who faced barriers to participation. Finally, although the REDAIM and TDF frameworks provide structured guidance, they cannot fully capture the complexities of communityDbased interventions, including informal practices, local power dynamics, and nuanced sociocultural influences on healthDseeking behaviour.

## Conclusion

The COMMIT programme strengthened communityDbased TB screening, diagnosis, and prevention in Cambodia. It improved case detection and treatment support by working closely with local authorities, health workers, and affected communities. The programme also demonstrated strong community engagement and sustained capacity-building. To maintain these gains, Cambodia will need stable financing, continued investment in community systems, and improvements in diagnostic and prevention strategies. Ending lowDvalue practices and adopting more efficient, personDcentred approaches will also be important. Overall, COMMIT demonstrates that communityDdriven, peopleDcentred models can make a lasting impact and can guide future TB efforts in Cambodia.

## Declarations

### Ethics approval and consent to participate

The National Ethics Committee for Health Research in Cambodia (NECHR) (reference: 430/NECHR) approved the study. A waiver of consent was approved for the secondary analysis of existing databases. Written informed consent was obtained from all interview participants.

Consent for publication Not applicable

### Availability of data and materials

The anonymised data may be available from the corresponding author upon reasonable request, subject to approval by the Ethics Committee.

### Competing interests

The authors declare no competing interests.

### Funding

The COMMIT programme and this evaluation were supported by funding from the USAID. The funders had no role in study design, data collection and analysis, decision to publish, or preparation of the manuscript.

### Authors’ contributions

AKJT conceptualised the study. AKJT, BS, MN-H, and SY developed the methodology. Data collection and curation were conducted by CP, HT, SM, SO, STep, and STuot. AKJT, BS, MN-H, and HT performed the formal analysis. AKJT, CP, SM, SCC, and STuot administered the project. Funding was acquired by AKJT, SCC, and STuot. AKJT, SY, and STuot provided overall supervision for the study. AKJT, BS, SE, and KMA drafted the manuscript. All authors reviewed and approved the final version.

## Acknowledgements

We acknowledge the contributions of the people and communities affected by TB to COMMIT, as well as the difficulties they have experienced as a consequence of the disease. We extend our gratitude to the COMMIT project staff, healthcare workers, community volunteers, and key partners (Khmer HIV/AIDS NGO Alliance (KHANA), Cambodian Health Committee (CHC), Health and Social Development (HSD), and Cambodia Anti-Tuberculosis Association (CATA)) for their dedicated efforts in implementing the programme. We also thank the National Centre for Tuberculosis and Leprosy Control (CENAT) for their invaluable support and collaboration. We thank the study participants for their time and qualitative insights.

## Notes

### Competing Interest Statement

The authors have declared no competing interest.

